# Physical activity specifically evokes release of cell-free DNA from granulocytes thereby affecting liquid biopsy

**DOI:** 10.1101/2021.09.01.21262910

**Authors:** Elmo W.I. Neuberger, Stephanie Sontag, Alexandra Brahmer, Keito F.A. Philippi, Markus P. Radsak, Wolfgang Wagner, Perikles Simon

**Affiliations:** Department of Sports Medicine, Rehabilitation and Disease Prevention, University of Mainz, Mainz, Germany; Helmholtz-Institute for Biomedical Engineering, Stem Cell Biology and Cellular Engineering, RWTH Aachen University Medical School, Aachen, Germany; Institute for Biomedical Engineering – Cell Biology, University Hospital of RWTH Aachen, Aachen, Germany; Department of Medicine III, Johannes Gutenberg University Medical Center, Mainz, Germany

## Abstract

Cell-free DNA (cfDNA) methylation-based diagnostics is a promising approach in oncology and hematooncology. Exercise impacts immune homeostasis and leads to a rapid and marked increase of cfDNA levels in blood. Since the origin of cfDNA during exercise remains elusive, the implications for liquid biopsy are unknown. In this study, we identified the source of cfDNA in 10 healthy untrained individuals before, immediately after, and 30 min after exercise, and in 6 patients with myeloid neoplasms or acute leukemia under resting conditions. A pyrosequencing assay was used to analyze the methylation levels of four CpGs, representing DNA from granulocytes, lymphocytes, monocytes, and non-hematopoietic cells. After exercise, cfDNA was almost exclusively released from granulocytes, with cell type specific proportions increasing significantly from 54.1% to 90.2%. Exercise did not trigger the release of cfDNA from lymphocytes or other analyzed cell types, whereas a small amount of cfDNA was released from monocytes. Compared to healthy people, patients with hematological malignancies show significantly higher cfDNA levels at rest with 48.1 (19.1; 78) vs. 8.5 (8.2; 9.5) ng/ml, data expressed as median (25th; 75th percentiles), and considerably higher levels of lymphocyte specific hypomethylated cg17587997 (*P*<.001). Hence, exercise-induced cfDNA elevations can compromise diagnostic accuracy.

**Key Points:**

- cfDNA is a robust sample source for targeted bisulfite sequencing, enabling reliable mapping of the source cells.
- cfDNA methylation signatures differ between healthy people and patients with hematological malignancies.
- During intense exercise, cfDNA is almost exclusively derived from granulocytes, which can affect results of liquid biopsy.

## Introduction

Circulating cell-free DNA (cfDNA) opened up a new horizon for genomic analyses, including absolute quantification, fragmentation profile analysis, detection of mutations and copy number variations, as well as epigenetic profiling.^1^ Cancers, including hematological malignancies, are accompanied by increased levels of cfDNA.^2,3^ More recently, research indicates that DNA methylation analyses are promising tools for diagnostic and prognostic purposes, and cfDNA can be utilized for studying global methylation profiles.^4,5^ Exercise impacts immune homeostasis and leads to rapid and markedly increased levels of cfDNA in blood. Depending on exercise modality, duration, and intensity cfDNA increases 2-20 fold,^6–9^ and even submaximal levels leads to 2-4 fold increases,^10^ showing a half-life of ∼15 min.^6,9^ Exercise-induced cfDNA releases could interfere with, or improve the diagnostic accuracy of methylation specific testing, depending on whether the DNA is derived from the clinically relevant cell type. However, the origin of cfDNA during exercise has not been studied in detail.

In a sex-mismatch transplantation model we already showed that the major part of cfDNA is released from cells of the hematopoietic lineage during exercise.^11^ Since the occurrence of neutrophil extra cellular traps (NETs) has been described following physical exhaustion,^12^ and cfDNA levels are correlated with markers of neutrophil activation including neutrophil elastase, and/or myeloperoxidase,^12,13^ it can be considered that granulocytes contribute to the pool of cfDNA.^14^ However, a more comprehensive picture is pending.

Schmidt et al. have shown that a targeted approach can be utilized to determine the cell types of origin reliably.^15^ In the present study, we use targeted methylation analysis with pre-validated CpG sites to assess the origin of cfDNA in healthy participants during exercise and patients with hematological malignancies under resting conditions. We provide evidence that targeted analysis of individual CpGs facilitates reliable, and fast analysis with low amount of starting material, and that exercise can interfere with clinical accuracy of methylation-based analysis.

## Methods

The study comprised of 10 healthy participants with a mean age of 26.0 ± 5.6 years and 6 patients aged 57.4 ± 11.6 with hematologic malignancies, including low risk myelodysplastic syndrome (LR MDS), chronic myelomonocytic leukemia (CMML), myelodysplastic/myeloproliferative neoplasm (MDS/MPN), myelofibrosis, and two patients with polycythemia vera. All diagnoses were confirmed by a hematopathologist according to the 2016 WHO criteria.^16^ The participants gave their informed consent to participate. All experimental procedures were approved by the Human Ethics Committee Rhineland-Palatinate and conformed to the standards of the Declaration of Helsinki of the World Medical Association.

### Exercise testing of healthy participants

The 10 healthy participants conducted an incremental running test on a treadmill, starting at 4 km/h. The speed was increased every three min by 1.5 km/h until volitional exhaustion as described by Ochmann et al.^17^ Venous blood samples were collected before (Pre), immediately after (Post), and 30 min after the test (+30’).

### Sample preparation, DNA extraction and bisulfite conversion

Whole blood samples were collected in tripotassium-EDTA covered Monovettes® (Sarstedt) and were processed within < 3 hours after sampling. Whole blood was centrifuged at 2,500 × g for 15 min. Separated plasma was centrifuged a second round at 2,500 × g for 15 min and stored at -80°C before further processing. DNA was extracted from 4 ml of plasma using QIAamp Circulating Nucleic Acid Kit (Qiagen) and eluted in 55 µl of UltraPure(tm) DNase/RNase-Free Distilled Water (Invitrogen). 50 µl of the eluate was bisulfite converted using the EZ DNA Methylation Kit (Zymo Research).

### Selection of cell type specific CpGs and deconvolution

Deconvolution of cell types is based on CG dinucleotides (CpGs) that are specifically hypomethylated in different cell types. Pre-validated CpG methylation sites were selected to estimate the origin of cfDNA from lymphocytes (cg17587997, FYN protooncogene (*FYN*)),^18^ monocytes (cg10480329, centromere protein A (*CENPA*)),^18^ and granulocytes (cg05398700, WD repeat domain 20 (*WDR20*)),^18^ and to differentiate leukocytes from other cells (cg10673833, myosin IG (*MYO1G*)),^15^ including endothelial cells, epithelial cells, fibroblasts, mesenchymal stem cells, hepatocytes, and muscle cells, described in Schmidt et al., 2020.^15^ We then generated a reference-based non-negative least-squares (NNLS) algorithm for the four CpGs of these cellular categories.^15,18^ An Excel calculation tool for cell type deconvolution based on the pyrosequencing measurements will be provided as a supplemental file after final publication of the article.

### Quantification of cfDNA and pyrosequencing

The cfDNA concentration was measured applying a pre-validated qPCR assay described in Neuberger et al., 2021.^19^ For pyrosequencing 4 µl of the bisulfite converted DNA were amplified with region-specific biotinylated/unmodified primer pairs (see supplemental file) using the PyroMark PCR kit (Qiagen) according to the manufacturer’s instructions using the following protocol: Initial activation at 95 °C for 15 min, 45 cycles of 30 s at 94 °C, 30 s at 56 °C, and 30 s at 72 °C followed by a final extension at 72 °C for 10 min. Pyrosequencing was performed on the PyroMark Q96 ID with the respective reagents (Qiagen).

### Data analysis and statistics

Statistical analyses were conducted with R version 4.0.2, using tidyverse version 1.3.0, and rstatix version 0.6.0. Graphical illustrations were prepared with ggplot2 version 3.3.2 and corrplot package version 0.85. Continuous data were log 10 transformed and tested for normal distribution with Shapiro-Wilk test. Non-normal distributed data were expressed as median (25^th^; 75^th^ percentiles). Global significant differences were tested with Friedman rank sum test. Paired or unpaired Wilcoxon rank sum tests with Bonferroni corrections, were used to compare within and between group differences.

## Results and Discussion

This study expands the knowledge about the origin of cfDNA during exercise and provides evidence that physical activity could compromise the diagnostic accuracy of epigenetic testing. Similar to other studies,^20,21^ we identified significantly increased absolute cfDNA concentrations in patients with myeloid neoplasms or acute leukemia (Figure A). During the all-out exercise test, which lasted 20.1 ± 5.4 min (mean ± SD), cfDNA increased ∼10 fold from 8.5 (8.2; 9.5) ng/ml to 80 (55.6; 142.7), decreasing to 19.8 (14.9; 28.9) +30’, with a typical half-life of ∼15 min.^22^

**Figure.**
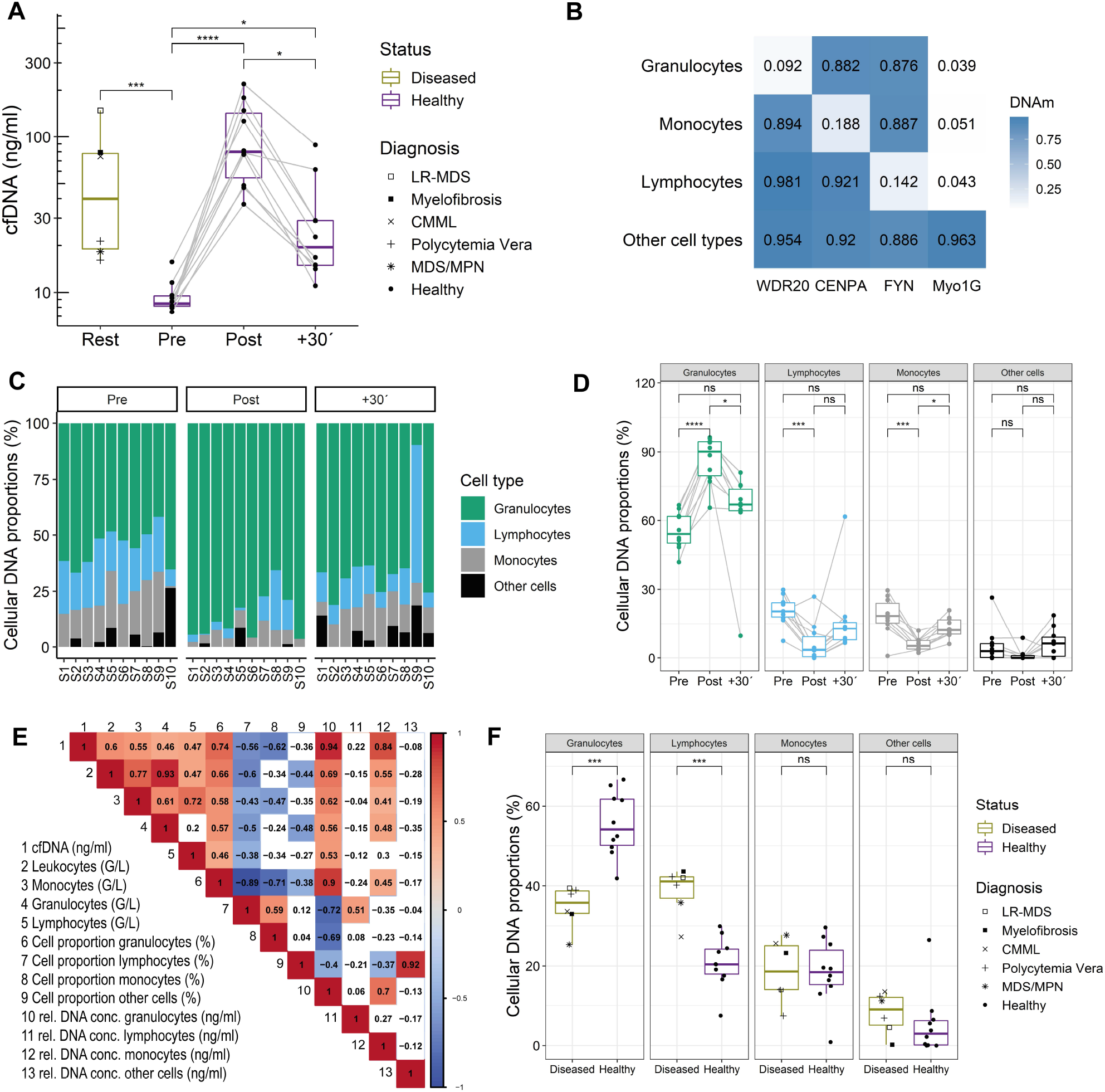
Concentration and origin of cfDNA in healthy persons during exercise and patients with hematological malignancies. (A) cfDNA concentration in blood plasma. (B) Heatmap of the percentage of methylation of the given CpGs in the reference matrix. (C, D) Deconvolution results indicating the origin of cfDNA samples before and after exercise in healthy subjects. (E) Spearman correlation matrix between cfDNA, blood counts, and deconvolution results. (F) Differences in cell type specific cfDNA proportions between healthy and diseased persons at rest. In all analysis *P*<.05 was considered significant. \**P*<.05, \*\**P*<.01, \*\*\**P*<.001, \*\*\*\**P*<.0001, ns: not significant.

To estimate the cellular composition, a non-negative least squares (NNLS) deconvolution algorithm was used, referring to the mean DNA methylation levels of the selected CpGs from different cell types (Figure B). As shown by Schmidt et al., the approach allows reliable estimation for the cellular composition by targeted analysis of the individual CpGs.^15^ Granulocytes are the major source of cfDNA during exercise. Cell type specific proportions increase significantly from 54.1% (50.1; 61.8) to 90.2% (79.7; 94.4) after exercise (Figure C,D), which is not reflected by the correlation of the cell count and cfDNA concentrations (Figure E). Monocyte and lymphocyte specific DNA proportions decreased significantly, but the relative DNA concentration, which was calculated by multiplying the cell type proportion with the cfDNA concentration, as measured by qPCR, indicated that a minor but significant proportion seems to be released from monocytes (Table). During acute exercise and 30 min after exhaustion, no DNA was released from other cell types including muscle cells and endothelial cells. A fast activation of granulocytes leading to vital or non-vital NETosis may cause cfDNA release during exercise.^23^

**Table.** Hematological response following exercise. Data are expressed as median (25th, 75th percentiles). The relative concentration (rel. conc.) of cfDNA from different cell types was calculated by multiplying the cfDNA concentration with the cellular DNA amount (%). Global statistical differences were calculated with non-parametric Friedman rank sum test.

|  | Pre | Post | +30' | Chi-Square | *P* |
| --- | --- | --- | --- | --- | --- |
| cfDNA (ng/ml) | 8.5 (8.2; 9.5) | 80 (55.6; 142.7) | 19.8 (14.9; 28.9) | 20 | 4,54E-05 |
| Leukocytes (G/L) | 6 (5.6; 6.1) | 9.5 (8.7; 12) | 7.1 (6; 7.5) | 15,8 | 0,000371 |
| Monocytes (G/L) | 0.4 (0.4; 0.6) | 0.7 (0.6; 0.8) | 0.5 (0.4; 0.6) | 15,2 | 0,0005 |
| Granulocytes (G/L) | 3.7 (3.1; 4.3) | 5.4 (4.4; 8.1) | 5.1 (3.5; 5.4) | 16,2 | 0,000304 |
| Lymphocytes (G/L) | 1.7 (1.4; 1.9) | 3.1 (2.4; 3.7) | 1.5 (1.3; 1.5) | 18,2 | 0,000112 |
| Cellular DNA monocytes (%) | 18.5 (15.3; 23.9) | 5.2 (3.8; 7.6) | 12.2 (10.4; 16.7) | 14,6 | 0,000676 |
| Cellular DNA granulocytes (%) | 54.1 (50.1; 61.8) | 90.2 (79.7; 94.4) | 66.9 (64.2; 73.8) | 18,2 | 0,000112 |
| Cellular DNA lymphocytes (%) | 20.4 (18; 24.2) | 3.5 (0.6; 9.2) | 12.9 (7.9; 15.5) | 11,4 | 0,003346 |
| Cellular DNA other cell types (%) | 3 (0.2; 6.2) | 0 (0; 1) | 6.3 (0.7; 9) | 2,6 | 0,272532 |
| Rel. conc. Monocytes (ng) | 1.7 (1.4; 2.2) | 4.9 (3.6; 6.9) | 3 (2.3; 4.7) | 9,8 | 0,007447 |
| Rel. conc. Granulocytes (ng) | 5.3 (4.4; 5.4) | 67.6 (46.7; 136.1) | 10.8 (9.3; 19.8) | 20 | 4,54E-05 |
| Rel. conc. lymphocytes (ng) | 1.9 (1.5; 2.3) | 2.1 (0.5; 8) | 2.1 (1.9; 3.7) | 0,6 | 0,740818 |
| Rel. conc. other cells (ng) | 0.3 (0; 0.7) | 0 (0; 0.8) | 1.2 (0.1; 1.6) | 1,4 | 0,496585 |

Under resting conditions, healthy and diseased persons show similar cfDNA proportions from monocytes and other cell types, whereas the levels of granulocyte and lymphocyte specific DNA differ significantly (Figure F, Table). The cfDNA levels of lymphocytes specific, hypomethylated cg17587997, which occur in the 5’UTR of FYN, are markedly increased and may be linked to pathogenesis.^24^ Since epigenetic changes crucially contribute to hematological malignancies,^25^ further analyses of relevant methylation sites have a great potential for monitoring disease burden and treatment response. Here we provide evidence that cfDNA can be used as a robust, non-invasive, and cost-efficient sample source for targeted sequencing approaches. Healthy and diseased persons show different methylation pattern of lymphocyte specific DNA. Exercise increases granulocyte specific DNA, which can affect diagnostic accuracy if resting periods are too short.

## Data Availability

All data are available from the corresponding author on reasonable request.

## Acknowledgements

The study was supported by intramural funding (Stufe 1) of the Johannes Gutenberg University Mainz and by the German Ministry of Education and Research (WW: VIP+, 03VP06120)

## Authorship Contributions

P.S., W.W., M.P.R., E.N. designed the research. K.F.A.P. and A.B. performed exercise testing and sample preparation. S.S., E.N. measured the samples and analyzed the data. E.N. drafted the manuscript with input from all authors. All authors read and approved the final manuscript.

